# Development of a Functional Needs Assessment Tool to estimate population level functional difficulties and need for services and assistive products

**DOI:** 10.64898/2026.07.10.26357317

**Authors:** Dorothy Boggs, Annet Birabwa, Suzanne Adkins, Oluwarantimi Atijosan-Ayodele, Sahan Bulathwela, Catherine de Cates, Allen Foster, Hannah Kuper, Cathy Holloway, Joseph Mugisha, Sarah Polack

## Abstract

**Background:** Globally, at least 2.6 billion people need rehabilitation services and more than 2.5 billion people need assistive technology (AT). However, reliable data are lacking on population level need for rehabilitation services and assistive products (AP) in different settings for evidence-based policy and programme planning. This first study paper describes the development of the Functional Needs Assessment Tool (FNAT), a new survey tool developed to fill this data gap between 2018 and 2023.

**Objective:** To develop a new multidomain tool to assess population-level functional difficulties and need for service and AP utilising both self-report and clinical assessment methodologies.

**Development stages:** FNAT was developed based upon primary and secondary data analysis, existing survey tools and expert consultation through a series of four steps: Step 1 Inform, Step 2 Build, Step 3 Draft and Step 4 Develop. FNAT uses both self-reported and clinical assessment tools to estimate the prevalence of functional difficulties/impairment and the need for services and AP in the following seven domains: vision, hearing, mobility, communication, cognition, self-care and mental health. It uses a two-stage population-based assessment with data collection through a bespoke tablet-based mobile application and web-based platform.

**Discussion:** FNAT is a new multi-domain modular tool developed to address data gaps by estimating prevalence of functional difficulties and service/AP needs in a population. Potential advantages and disadvantages were highlighted during the development stages, and the tool needs to be pilot tested to assess the feasibility of the methodology and the functionality of the tablet-based mobile data collection application.

## 1. INTRODUCTION

Rehabilitation and assistive technology (AT) can be critical for promoting independence, participation and well-being among people with functioning difficulties.^1, 2^ Yet, globally, unmet need for these essential services remains high; at least 2.6 billion people experience difficulties with functioning and could benefit from rehabilitation ^3, 4^ and over 2.5 billion people need assistive technology (AT) ^5^. Reliable data on population need to inform policy and service provision are currently lacking, especially in low- and middle-income country (LMIC) settings where access to both rehabilitation and assistive products (AP) is often limited.^6^

The International Classification of Functioning, Disability and Health (ICF) defines functioning as an umbrella term for body functions, body structures, activities, and participation.^7^ It denotes the interaction between an individual (with a health condition) and their contextual (environmental and personal) factors (see **Supplementary File 1**).^7^ Alongside mortality and morbidity, functioning is increasingly recognized as the third health indicator, yet data to measure and monitor it are lacking.^8–10^ This is due, in part, to the challenge of reliable identification of functioning and related service/AP need within surveys.

Functioning and service/AP need can be assessed through self-report. Examples include Washington Group (WG) questions on functioning, widely used in surveys of disability, which assess self-reported difficulty (“none”, “some”, “a lot” or “cannot do”) in domains of vision, hearing, mobility, cognition, self-care, communication and mental health ^11^. The WHO rapid Assistive Technology Assessment (rATA) survey tool collects data on participant self-reported AP need.^12^ Self-report methods are relatively rapid, simple and capture participants’ own experience of their functioning and preferences. However, limitations include potential inaccuracy in reporting; for example, some people with impairments may be unaware that they need surgical/medical interventions rather than AP.^13–15^

Another approach involves clinicians assessing impairment severity, cause, diagnosis and related need for services/AP. This can provide a more accurate assessment of service/AP needs ^16^, but can be time-consuming and requires trained clinicians. “Rapid” clinical impairment assessment survey methodologies often integrate mobile health technology to overcome some disadvantages of traditional clinician-led approaches. For example, the Rapid Assessment of Hearing Loss (RAHL) ^17^ uses the mobile audiometry tool hearTest ^18^ together with a clinical ear examination; and the Rapid Assessment of Avoidable Blindness (RAAB) ^19^ uses the mobile tool Peek Vision ^20–22^ together with a simplified ophthalmic examination. However, these approaches often focus on a single domain only (e.g. vision) and typically lack assessment of the individual’s functioning, environment and preferences, which are important for determining appropriate service/AP need.^8, 13, 14^ Comprehensive survey methods which incorporate self-report, clinical impairment and functional assessment of service/AP need are lacking.

In response to these gaps and challenges, the Functional Needs Assessment Tool (FNAT), a new multi-domain survey methodology and bespoke mobile data collection application, was developed. FNAT aims to estimate the i) prevalence of functional difficulties/impairment and ii) need for related health and rehabilitation services and AP. In this paper we describe the development of FNAT.

## 2. DEVELOPMENT STAGES

FNAT’s development included four steps completed between 2018-2023: 1) INFORM assessment methods through data analysis; 2) BUILD using widely used international disability and functional domain-specific assessment tools; 3) DRAFT a functional needs assessment to identify service/AP need; and 4) DEVELOP a bespoke tablet-based mobile application and web-based platform for data collection. As each step built on the previous one, the methods and results are presented sequentially for each.

FNAT focuses on the functional domains of vision, hearing, mobility, communication, cognition, self-care and mental health, selected to align broadly with WHO Global Cooperation on Assistive Technology (GATE)’s AP subgroupings ^1, 23^ and ensure a holistic focus on functioning ^7^. In this paper, “service need” refers to need for health and rehabilitation services.

### Step 1: INFORM

Primary and secondary data analysis from previous population-based surveys in the domains of vision, hearing, mobility and cognition, informed the FNAT methods for assessing service/AP need. These analyses have been published elsewhere ^13, 14, 16, 24–27^. In brief, our analyses found that self-report alone can both over and under-estimate service/AP need and that clinical impairment assessment is more accurate but doesn’t capture individual’s functioning, environment and preferences.^8, 13, 14^ These findings supported the development of a hybrid approach combining self-report, clinical impairment and functional assessments.^13, 14^

Clinician-led assessments, on all survey participants, across multiple domains are time consuming and costly, which in turn, may limit wide survey implementation.^13, 14, 24–28^ Therefore, survey data were analysed to explore the use of a two-stage assessment approach, involving rapid “Stage 1” screening by non-clinicians followed by clinician-led assessments for those screening positive. Our analyses explored the use of the WG questions as first-stage screening tool followed by clinician led assessment for people who report any difficulty in one domain.^16, 28^ This two-stage approach was found to identify i) most people with disabilities if “some/worse difficulty” was reported in a specific domain and ii) most people with service/AP need, but only with moderate sensitivity and specificity. The later finding highlighted a need to include additional brief first-stage tools (described below) into FNAT to improve sensitivity and specificity.

### Step 2: BUILD

In this step, the two-stage assessment was developed through incorporating or adapting existing International Centre for Evidence in Disability (ICED) survey tools and expert consultation.

#### Stage-one assessment

To improve “screening” sensitivity, the following rapid, domain-specific self-report tools were identified for administration by non-clinicians, alongside the WG Extended Question set (18+ years) and WG/UNICEF Child functioning questions (children 2-18 years) ^11^: i) six validated screening questions used in Rapid Assessment of Musculoskeletal Impairment (RAM) ^29^, ii) three questions from a Communication Disability Screening Tool ^30, 31^ (CDST, see more details below), and iii) incontinence questions modified from the ICED Vanuatu Women, Water and Disability Study ^32^ (see **Box 1**). In addition, validated rapid-mobile impairment screening tools widely used in surveys for vision (e.g. Peek Vision ^20–22^) and hearing (e.g. hearTest ^18^), were also considered for first-stage assessment and need to be pilot tested.

**Box 1.**
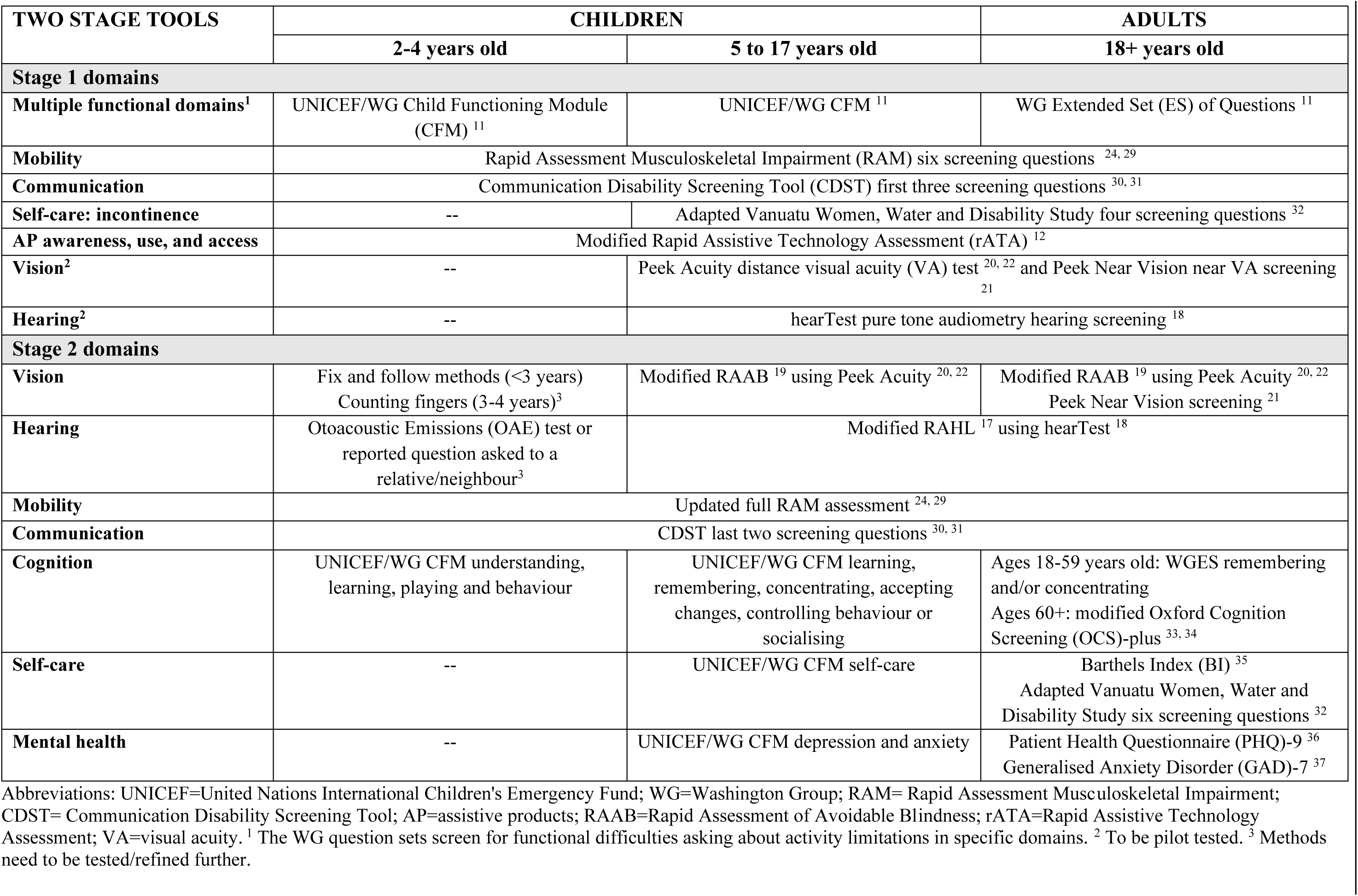
The two-stage functional difficulty/impairment screening and assessment tools/questions selected for FNAT by domain and age groups.

Domain specific criteria were developed for “screening positive” at stage 1 and requiring stage 2 clinician assessment, informed by previous research ^13, 14, 16, 24–27^. These are shown in **Table 1** and need to be pilot tested.

**Table 1.**
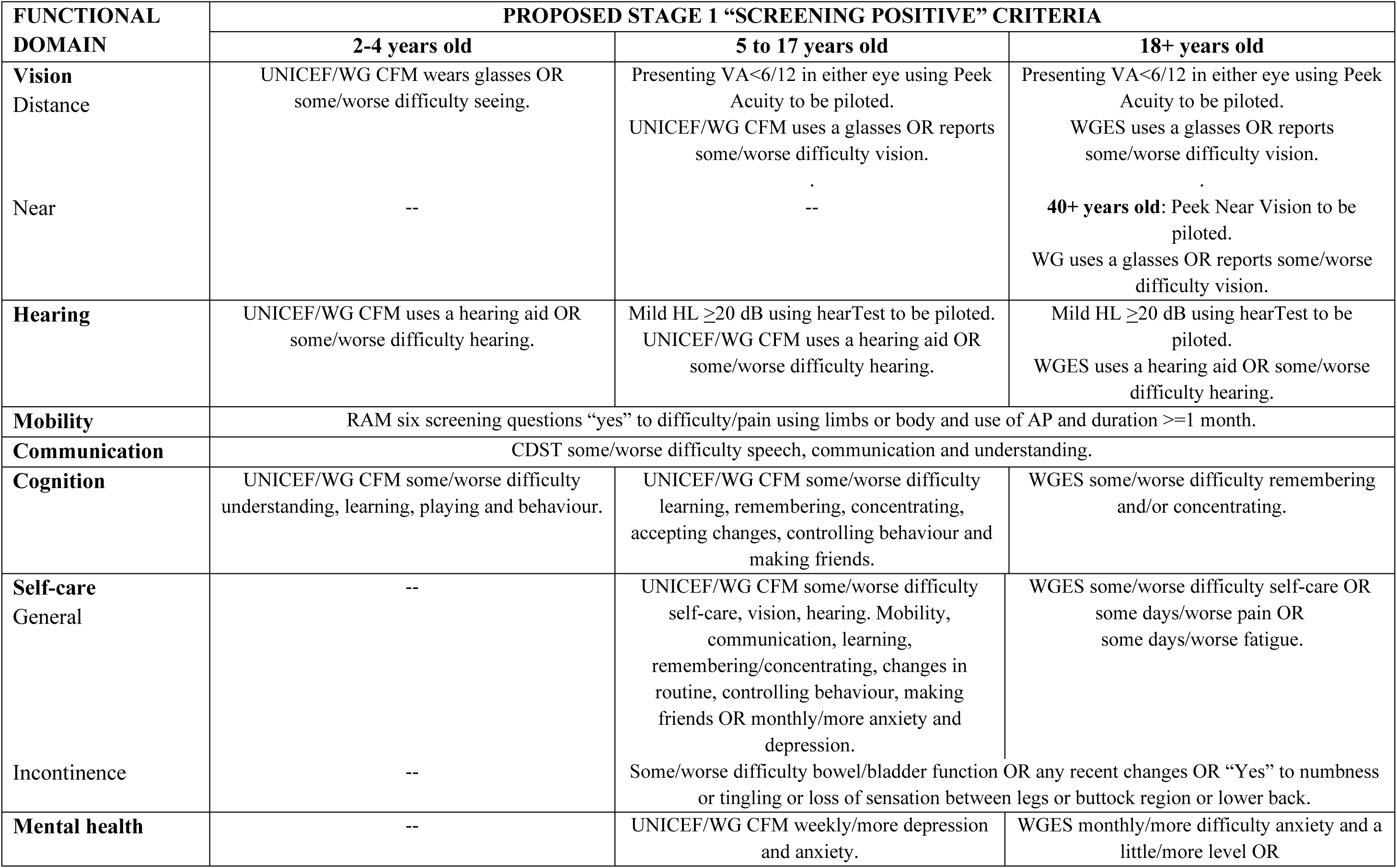
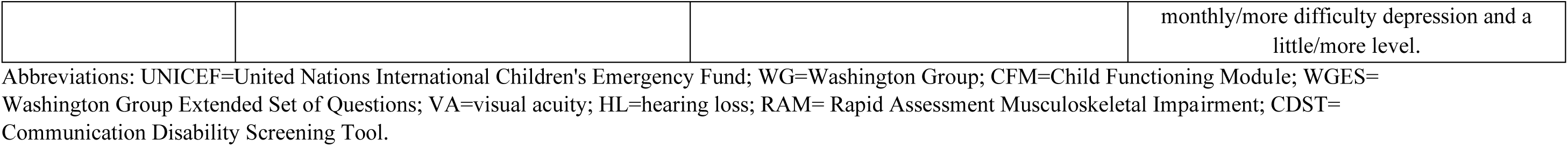
Proposed criteria for “screening positive” at Stage 1 by age and domain to be pilot tested.

#### Stage-two clinician assessment

The seven functioning domains require different assessment approaches and availability of validated clinical assessment tools suitable for surveys varies by domain (See **Box 1** and **Figure 1**).

**Figure 1.**
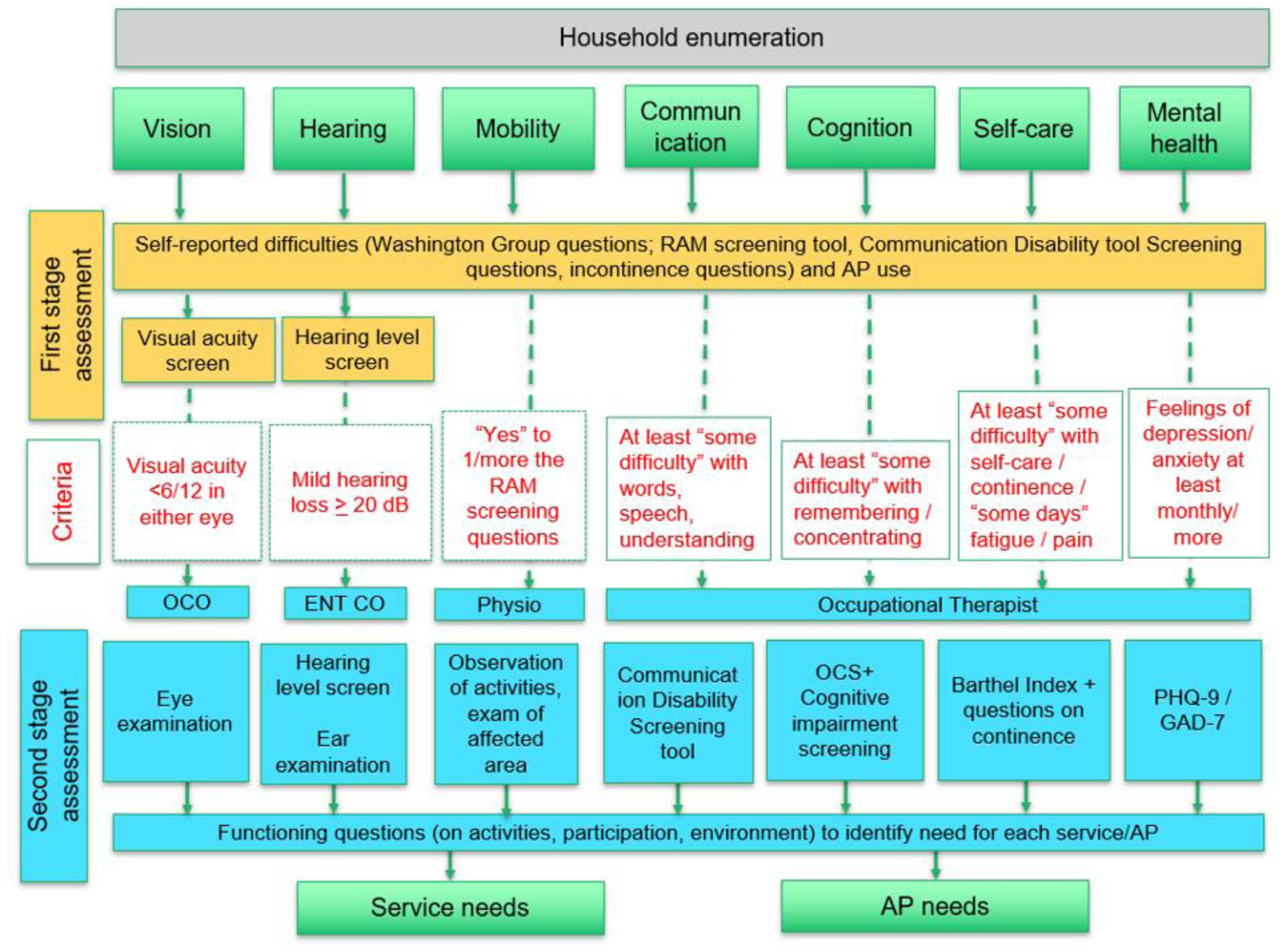
Overview of data procedures of the Functional Needs Assessment Tool (FNAT).

For FNAT vision, hearing and mobility modules, we drew on methods from existing single domain rapid survey methods including RAAB ^19^, RAHL ^17^ and RAM ^24, 29^. Briefly, these involve standardised, simplified assessments administered by a relevant clinician to determine the presence, severity and likely cause of impairment which informs service/AP need. These methods are published elsewhere ^17, 19, 24, 29^.

For self-care, Barthels Index (BI) ^35^, a tool to assess activities of daily living in clinics and surveys that is widely used internationally ^35, 38–40^, was selected for participants aged over 17 years. Additional questions on continence were adapted from the Vanuatu Women, Water and Disability Study.^32^

For mental health, two tools validated in different LMICs ^41–43^ were included (for participants aged 18+): the Patient Health Questionnaire (PHQ)-9 ^36^ for reported depression, and the Generalised Anxiety Disorder (GAD)-7 ^37^ item tool.

For communication and cognition, rapid assessment tools for use in population surveys where literacy levels are low are lacking.^44, 45^ A Communication Disability Screening Tool (CDST) was identified through expert consultation. This rapid, simple tool was co-developed by speech and language therapists and the United Nations High Commissioner for Refugees (UNHCR) for use with the refugee population in Rwanda.^30, 31^ It includes three screening questions about the level of difficulty with speech, communication and understanding, followed by questions on the impact and nature of communication disability.

For cognition, the Oxford Cognition Screening (OCS)-plus, developed by Oxford University, is a domain-general cognitive impairment screening tool administered using a pre-programmed computerised tablet-based tool that has been validated, including with populations with low-literacy.^33, 34^ This tool is relatively time-consuming to administer (approximately 20-25 minutes) therefore the feasibility of including this needed testing.

### Step 3: DRAFT

#### Services and assistive products

FNAT aims to assess need for services and AP. We compiled a core set of common i) health and rehabilitation services and ii) AP (see **Supplemental File 2**) based on previous surveys ^13, 14, 16, 17, 19, 24–27^, WHO GATE AP lists ^1, 23^ and discussion with stakeholders. The list of service/AP use and need indicators can be adapted based on availability in different settings.

#### Functional assessment

In determining service/AP needs, a gap identified from previous surveys is a holistic functional assessment approach that takes into account all ICF components, such as activities, participation and context, as well as impairment/body function.^8, 13, 14, 16, 25, 46^ To address this gap, for FNAT, we developed functional assessment questions for each domain. Existing general tools, such as Model Disability Survey personal support questions ^47^, and domain specific tools, such as the six-part scale from the Functional Vision questionnaire ^48^, were referenced, mapped and compiled. For each AP (**Supplemental Table 2B**), a short set of questions (2-6) were developed focusing on relevant activities, participation, contexts and preference. Questions for these “AP decision trees” were also informed from the lead author’s Occupational Therapy clinician background knowledge and by AP knowledge rules developed by experts for a WHO project “Assistive Product Explorer (ASPREX)” ^49, 50^. Prior to pilot testing, the questions were iteratively developed including feedback from 1-2 clinicians per domain.

#### Determining service/AP need

Throughout each domain module, self-report, clinical impairment and functional assessment data were integrated to guide the clinician to assign service/AP intervention needs. This was implemented through questionnaire programming using i) skip features and ii) section-level and question-level screening or “filter” criteria within each module (see **Figure 1** and **Supplemental File 2**). For some interventions, this was more standardised: for example, a person identified clinically as having uncorrected refractive error or near vision impairment would be assigned as needing distance glasses or near vision glasses respectively ^19^. For others, a combination of information and clinical judgement would be required: for example, for a person identified as having moderate mobility impairment, their answers to specific functional assessment questions (e.g, about family, home and community participation, activities and environments) alongside clinical reasoning, would be used to assess whether a wheelchair and/or walking aid was needed. It will be important to assess the feasibility and accuracy of these additional questions to provide improved information for the clinicians’ assessment of service/AP need. Additionally, for five priority AP (distance and near vision glasses, hearing aids, wheelchairs and prosthetics), “AP need decision trees” integrating assessment methods were developed and need to be pilot tested.

### Step 4: DEVELOP

A bespoke data collection software application (Android-based for tablets) and web-based platform (“mFNAT”) were developed to integrate the FNAT methodology including linking to the three mobile impairment screening tools (e.g. Peek Vision, hearTest and OCS-Plus) and displaying WHO AP images ^51^. mFNAT was developed to include i) multi-lingual translation capabilities, ii) time-stamped and device-stamped data, iii) ability to collect data offline and synchronise data up/down once online, iv) user permission rights for different data collector roles, and v) a web-based dashboard for data and quality monitoring. It also included i) section and question-specific skips/filters, ii) autofill features, and iii) systems for identifying and displaying people “screening positive” and requiring assessment by a clinician. Codebooks were developed to link variables when relevant across the functional domain modules.

The development of mFNAT was undertaken by an app development consultant team based in Sri Lanka with Global Health for Development (GH4D) with input and support from the AT2030 consortium ^52^, University College of London (UCL) and London School of Hygiene and Tropical Medicine (LSHTM) (see **Figure 2**). The bespoke platform was set up on a dedicated AT2030 project host encrypted and password protected server at UCL.

**Figure 2.**
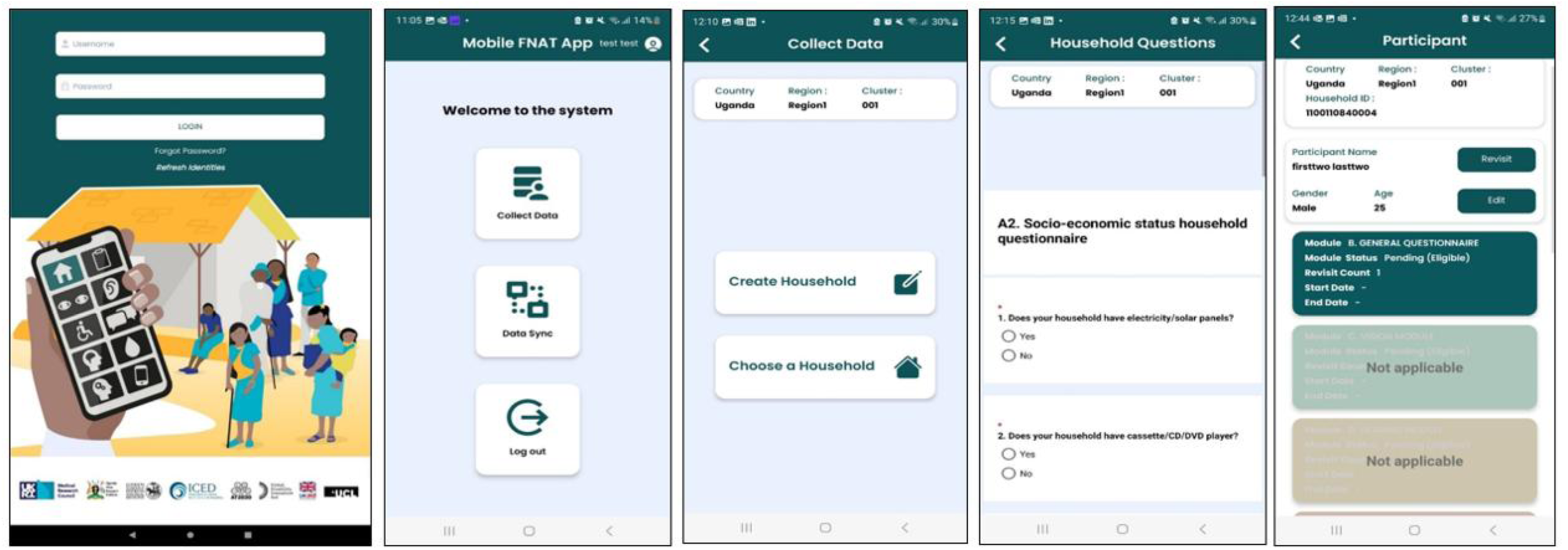
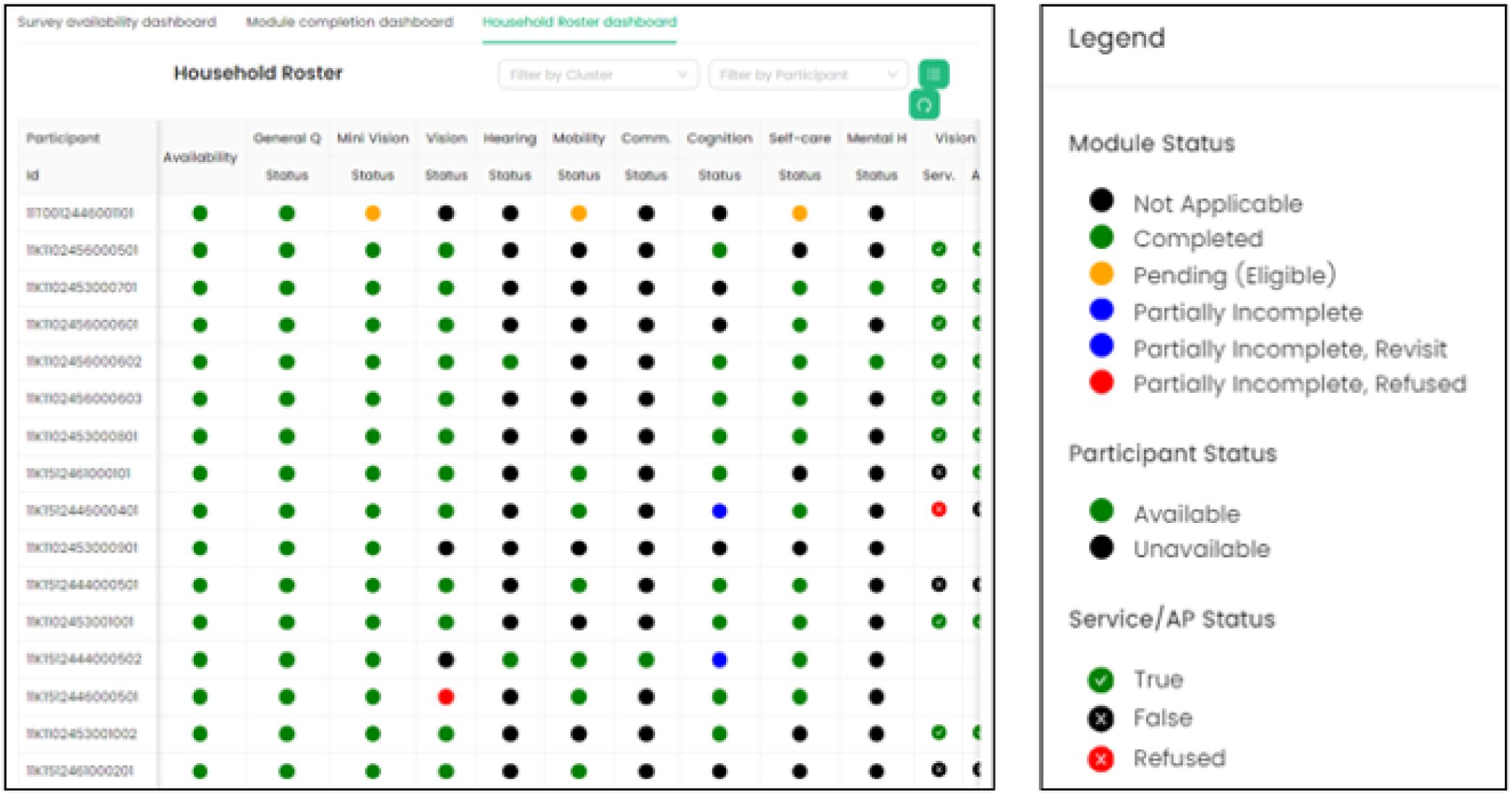
Screen shot examples of the mobile FNAT (mFNAT) **2A.** mFNAT tablet-based mobile application for data collection **2B.** mFNAT web-based platform for data collection.

## 3. DISCUSSION

FNAT is a novel multidomain population-based survey tool to estimate functional difficulties/impairments and need for services and AP, integrating self-report, clinical impairment and functional assessment methodology using a bespoke mFNAT modular tablet-based mobile application and web-based platform.

Throughout the development of the tool, potential advantages and disadvantages have been highlighted.^13, 14, 16, 24–27^ First, FNAT data will provide estimates of functional difficulties and service/AP need across seven domains which will be critical to gain a more holistic, reliable, evidence-based understanding of the population’s functional needs across the life course and address a number of cross-sectoral data gaps. Previous methodology utilised in population-based surveys is typically single domain focus, providing estimates for one impairment type ^17, 19^, or alternatively relies on self-report only for service/AP needs ^12, 47^ which has limitations. Second, FNAT uses a hybrid methodology integrating self-report and clinician assessment methodology. Third, FNAT’s methodology was built using a tablet-based mobile data collection application and web-based platform, which enabled the development of a multi-domain assessment tool integrating three separate mobile impairment screening tools ^18, 20, 33, 34^. Once tested, this software has potential to enhance survey team coordination, save time and resources, and provide advanced data monitoring and quality mechanisms through the dashboard.

However, there are some limitations to the FNAT methodology. First, the survey tool is relatively complex, and training, time and human resource intensive. It will need pilot testing in different settings to assess the feasibility of the methodology and the functionality of the tablet-based mobile data collection application, and refined and updated accordingly. Second, we have recommended specific clinical cadres for FNAT administration based upon previous surveys ^17, 19, 24, 29^, but the availability of clinicians, and specifically rehabilitation professionals, is limited in some settings and professional qualifications and responsibilities can vary considerably across countries.^53, 54^ Third, overall FNAT proposes to assess seven functional domains, 15 related services and 26 related AP, but these areas might not encompass the population’s actual needs. Though there are potential cost savings from combined domain assessment ^55^, the specific functional domains and services/AP assessed should be driven by the context, including data needs, and availability of resources and personnel.

In future, there could be areas for additional FNAT development. For example, there may be the potential to develop a “Rapid” version, which could include restricting to 40+ years old to collect the majority of data for service planning with a smaller sample size, and/or a “Short” version with a minimal set of questions required ^56^.^27^ Additionally, a cadre accuracy testing study ^57^ could be conducted to assess whether one clinician cadre, such as a physiatrist, could administer the full survey and/or if a non-clinical cadre, such as a community health worker, could administer a combined multidomain functional assessment section once more advanced service/AP decision trees are developed.^27^

## 4. CONCLUSION

FNAT is a new multi-domain survey tool for estimating prevalence of functional difficulties/impairments and service/AP needs in a population, and needs to be pilot tested in different settings. Given the SDG agenda to reach the most vulnerable, and the multidomain and multisectoral nature of functional need assessment, it is hoped that this tool will provide much needed data to provide functioning data across sectors for policy, programming and planning and strengthen the evidence-base to improve the imperative of “inclusive access for all”.

## Author Contributions

Conceptualization: D.B., S.P., H.K. and C.H.; Methodology: D.B., S.P., H.K., A.F., A.B., J.M., S.A., C.deC., O.A., and S.B.; writing—original draft: D.B., with input from S.P.; writing—review and editing: all authors; Supervision: S.P., H.K., A.F. and J.M.; Funding Acquisition: H.K., S.P., and C.H.; Software: D.B., S.P., S.B., C.H., S.A., and C.deC.; Resources and project administration: A.B. and D.B.; Visualisation, Data curation, Formal analysis: D.B. All authors have read and agreed to the published version of the manuscript.

## Data Availability

The original contributions presented in this study that informed tool development are included in the article/supplementary material, further inquires can be directed to the corresponding author.

https://datacompass.lshtm.ac.uk/id/eprint/3863/

## Acknowledgments

The authors wish to acknowledge the many collaborators involved in the funding and design of the FNAT methodology. We thank Peek Vision, hearX and OCS-Plus and colleagues for their support in the use of their screening apps and integration with mFNAT. Additionally we thank Julie Marshall and Helen Barrett for their communication tool guidance, and the EN-BIRTH Study Group for sharing the EN-BIRTH E-data app as an example for the mFNAT team. Finally, we thank colleagues from the International Centre for Evidence in Disability for their input into the survey protocol development, colleagues from UCL for their mFNAT project management support and the GH4D Sri Lankan-based consultant team for the mFNAT tablet-based mobile application and web-based platform development. We are grateful to AJ Simpson and Jyoti Shah for project administration and financial support.

## Supplementary Material

The following is available online, Supplementary file 1: FNAT terminology and definitions used in the paper and Supplemental File 2: Proposed service and assistive product need indicators clinically assessed in FNAT in each functional domain

