## Supplemental files for "Development of a Functional Needs Assessment Tool to estimate population level functional difficulties and need for services and assistive products"

**Supplementary file 1: FNAT terminology and definitions used in the paper**

| **TERM** | **DEFINITION** |
| --- | --- |
| Assistive products | Any external product (including devices, equipment, instruments or software), especially produced or generally available, the primary purpose of which is to maintain or improve an individual’s functioning and independence, and thereby promote their well-being. AP are also used to prevent impairments and secondary health conditions.^1, 2^ |
| Assistive technology | The application of organized knowledge and skills related to assistive products, including systems and services. ^1, 2^ |
| Functioning | An umbrella term in the International Classification of Functioning, Disability and Health (ICF) for body functions, body structures, activities, and participation (see **Figure** below). It denotes the positive aspects of the interaction between an individual (with a health condition) and that individual’s contextual factors (environmental and personal factors).^3^ |
| Rehabilitation | A set of interventions designed to optimise functioning and reduce disability in individuals with health conditions in interaction with their environment.^4^ |

**Figure.** International Classification of Functioning, Health and Disability diagram^3^
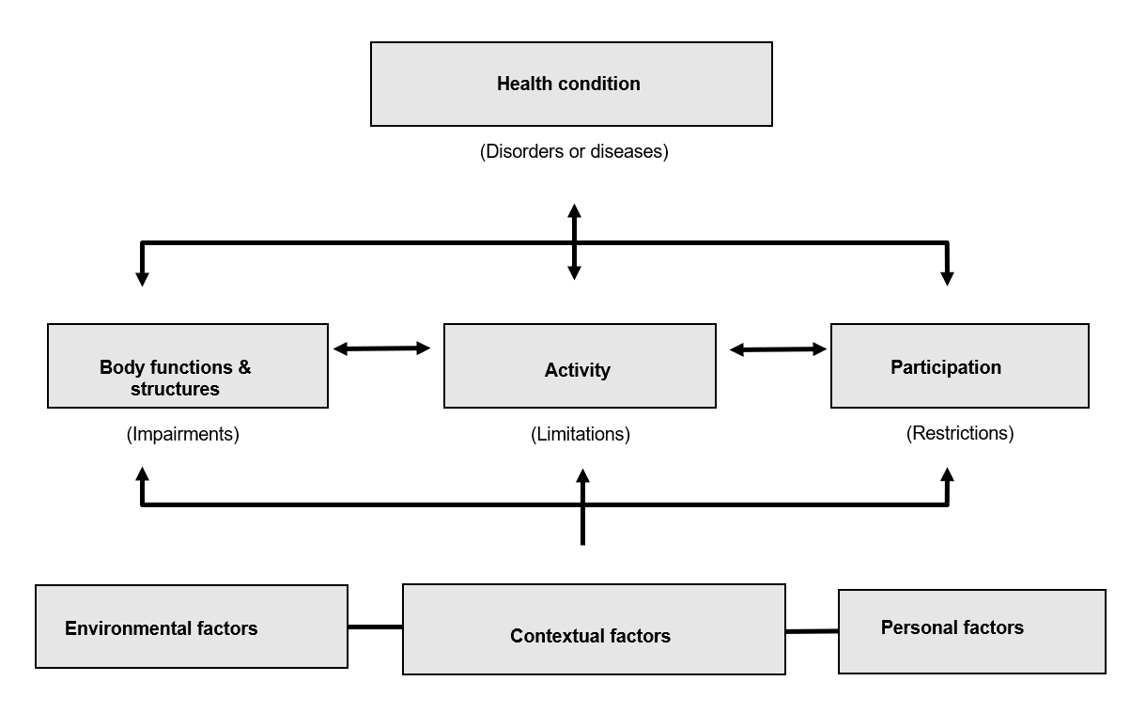

**Supplemental File 2:** Proposed service and assistive product need indicators clinically assessed in FNAT in each functional domain

**2A.** List of service use and unmet need indicators*

|  | **SERVICE UNMET NEED INDICATORS ASSESSED IN EACH DOMAIN** | | | | | | |
| --- | --- | --- | --- | --- | --- | --- | --- |
| **LIST OF SERVICE USE INDICATORS** | **VISION** | **HEARING** | **MOBILITY** | **COMMUNICATION** | **COGNITION** | **SELF-CARE** | **MENTAL HEALTH** |
| **Surgery** |  |  |  |  |  |  |  |
| **Medication** |  |  |  |  |  |  |  |
| **Eye examination** |  |  |  |  |  |  |  |
| **Low vision service** |  |  |  |  |  |  |  |
| **Vision rehabilitation** |  |  |  |  |  |  |  |
| **Hearing examination** |  |  |  |  |  |  |  |
| **Hearing rehabilitation** |  |  |  |  |  |  |  |
| **Physiotherapy** |  |  |  |  |  |  |  |
| **Occupational therapy** |  |  |  |  |  |  |  |
| **Speech therapy** |  |  |  |  |  |  |  |
| **Prosthetics and orthotics services** |  |  |  |  |  |  |  |
| **Counselling/psychosocial support** |  |  |  |  |  |  |  |
| **Information on exercises without ongoing rehabilitation** |  |  |  |  |  |  |  |
| **Other rehabilitation** |  |  |  |  |  |  |  |
| **Environmental modification** |  |  |  |  |  |  |  |

* If the box is shaded with a tick mark, the service need indicator is assessed in the functional domain. If the box is empty, the service need indicator is not assessed in the functional domain.

**2B.** List of assistive product use and unmet need indicators*

|  | **ASSISTIVE PRODUCT (AP) UNMET NEED INDICATORS ASSESSED IN EACH DOMAIN** | | | | | | |
| --- | --- | --- | --- | --- | --- | --- | --- |
| **LIST OF AP USE INDICATORS** | **VISION** | **HEARING** | **MOBILITY** | **COMMUNICATION** | **COGNITION** | **SELF-CARE** | **MENTAL HEALTH** |
| **Mobile phone/smart PDA/tablet** |  |  |  |  |  |  |  |
| **Long distance glasses** |  |  |  |  |  |  |  |
| **Reading glasses** |  |  |  |  |  |  |  |
| **Low vision glasses** |  |  |  |  |  |  |  |
| **Magnifying glasses, telescopes** |  |  |  |  |  |  |  |
| **White cane** |  |  |  |  |  |  |  |
| **Pill organisers** |  |  |  |  |  |  |  |
| **Audio players** |  |  |  |  |  |  |  |
| **Talking and touching watches** |  |  |  |  |  |  |  |
| **Alarm signallers** |  |  |  |  |  |  |  |
| **Hearing aids** |  |  |  |  |  |  |  |
| **Walking aids** |  |  |  |  |  |  |  |
| **Crutches** |  |  |  |  |  |  |  |
| **Stick, cane, tripod and quadripod** |  |  |  |  |  |  |  |
| **Walking frame or rollator** |  |  |  |  |  |  |  |
| **Wheelchairs** |  |  |  |  |  |  |  |
| **Prosthetics** |  |  |  |  |  |  |  |
| **Orthoses** |  |  |  |  |  |  |  |
| **Protective footwear** |  |  |  |  |  |  |  |
| **Toilet chair/commode** |  |  |  |  |  |  |  |
| **Shower/bath chair** |  |  |  |  |  |  |  |
| **Grab bars** |  |  |  |  |  |  |  |
| **Ramps** |  |  |  |  |  |  |  |
| **Continence products** |  |  |  |  |  |  |  |
| **White boards- simple memory supports** |  |  |  |  |  |  |  |
| **Communication boards or books** |  |  |  |  |  |  |  |

* If the box is shaded with a tick mark, the AP need indicator is assessed in the functional domain. If the box is empty, the AP need indicator is not assessed in the functional domain. Abbreviations: AP=assistive product/s.

1.	World Health Organization. Global report on assistive technology. 2022.

2.	World Health Organization. Priority assistive products list. Geneva: WHO. 2016.

3.	World Health Organization. International Classification of Functioning, Disability and Health (ICF) 2001 [Available from: <https://www.who.int/standards/classifications/international-classification-of-functioning-disability-and-health>.

4.	World Health Organization (WHO). Rehabilitation 2030: A call for action. Geneva, Switzerland: WHO.
